# Psychological factors and weight trajectory after bariatric surgery: the role of eating disorders, psychiatric conditions, and exposure to violence

**DOI:** 10.64898/2026.04.30.26351759

**Authors:** Claire Nominé-Criqui, Blanche Nitting, Pierrette Witkowski, Nicolas Reibel, Didier Quilliot, Coralie Gaspard, Laurent Brunaud

**Affiliations:** Nancy University Hospital, Department of Visceral, Metabolic, and Oncologic Surgery, Nancy, France; Medicine Faculty of Nancy, Lorraine University; Department of Endocrinology, Diabetology, Nutrition, Nancy University Hospital, Nancy, France

**Keywords:** Bariatric surgery, eating disorders, weight regain, obesity, weight trajectory

## Abstract

**Background:** Psychiatric conditions, eating disorders (EDs), and exposure to violence are highly prevalent in patients with severe obesity. However, their association with postoperative weight trajectories following bariatric surgery remains unclear.

**Objective:** To assess the associations between eating disorders, psychiatric conditions, and history of violence with multiple dimensions of weight trajectory after bariatric surgery.

**Methods:** This retrospective study included 414 patients with severe obesity from the OBESEPI cohort who underwent bariatric surgery at Nancy University Hospital (France). Psychological factors were assessed using a standardized preoperative psychiatric interview. Weight outcomes included preoperative BMI change, maximal BMI loss (ΔBMImax), final BMI change (dBMIdf), weight regain (BMIR), and magnitude of weight regain (dBMIR). Multivariable linear and logistic regression models were adjusted for age and sex.

**Results:** Psychological factors were not associated with baseline BMI or preoperative BMI variation. A history of violence was significantly associated with greater maximal BMI loss (β = 1.99, 95% CI [0.73–3.26]; p = 0.002) and greater final BMI reduction (β = 1.81, 95% CI [0.47– 3.14]; p = 0.009). Eating disorders and psychiatric conditions were not associated with weight loss outcomes. No association was observed between overall exposure to violence and weight regain. However, subtype analyses showed that physical violence was associated with a higher risk of weight regain, whereas psychological violence was associated with a lower risk. No significant associations were found for the magnitude of weight regain.

**Conclusions:** Eating disorders and psychiatric conditions were not associated with postoperative weight outcomes in this cohort. In contrast, exposure to violence—particularly when differentiated by subtype—was associated with distinct patterns of weight loss and regain. These findings highlight the relevance of trauma-informed assessment in bariatric care and support a more individualized approach to obesity management.

## Introduction

Obesity is a multifactorial condition involving complex psychological, behavioural and biological interactions, extending beyond metabolic dysregulation. Several studies have reported a higher prevalence of obesity among individuals with psychiatric disorders compared with the general population [1, 2]. This relationship is likely bidirectional. A meta-analysis by Luppino et al. showed that obesity increases the risk of depression, while depression also increases the risk of obesity [3].

Several mechanisms may explain this association. Neuroendocrine dysregulation, particularly involving the hypothalamic–pituitary–adrenal (HPA) axis, can lead to chronic hypercortisolism, increasing appetite and visceral fat accumulation. Alterations in reward-related neural circuits, especially within the dopaminergic system, may promote maladaptive eating behaviours such as compulsive food intake and emotional eating [4]. In addition, obesity is associated with a chronic low-grade inflammatory state, which may contribute to the development of mood disorders [5–7].

The gut microbiota–brain axis has recently been described as a key pathway linking metabolic and psychological processes. The gut microbiota is now recognised as an important regulator of metabolism, immune signalling and neuroendocrine function. Alterations in microbial composition and diversity have been observed in individuals with obesity and are thought to influence energy balance and metabolic homeostasis [8]. The gut microbiota can also interact with the central nervous system through neural, immune and metabolic pathways, contributing to the regulation of appetite, mood and behaviour [9, 10]. Dysbiosis has been associated with psychiatric disorders, particularly depression and anxiety, leading to the concept of “psychobiotics” [8]. Bariatric surgery induces significant changes in gut microbiota composition, which may partly mediate its metabolic effects [11, 12].

Among psychiatric factors, eating disorders (EDs) are particularly relevant. They are more prevalent in individuals with obesity, especially among candidates for bariatric surgery, where rates of binge eating disorder (BED) can reach 30–50% [13]. EDs may contribute to both the development of obesity and postoperative outcomes, particularly weight regain. Behavioural patterns such as loss of control eating and emotional dysregulation are likely to play a central role in this association.

Current clinical guidelines, including those from the French National Authority for Health (HAS, 2024) [14], recommend systematic psychological and psychiatric assessment prior to bariatric surgery to identify factors that may affect adherence, weight outcomes and long-term prognosis. However, the impact of psychiatric factors on weight evolution remains unclear, as most studies have focused on isolated outcomes such as total weight loss or weight regain. Weight trajectory after bariatric surgery is a dynamic process involving several phases.

Analysing weight trajectories using multiple indicators—such as maximal weight loss, long-term evolution and weight regain—may provide a more comprehensive understanding of postoperative outcomes.

In this context, we aimed to explore the association between eating disorders, psychiatric conditions, and a history of violence with different aspects of weight trajectory in patients undergoing bariatric care.

## Methods

### Study design and population

This study is a retrospective analysis based on data from the “Obésité sévère et épigénétique” (OBESEPI) prospective interventional cohort [15, 16]. The cohort was established between 2013 and 2017 and includes patients with severe obesity who underwent bariatric surgery and were followed at Nancy University Hospital (France).

A total of 454 patients were initially included in the cohort. Patients with missing data on key variables were excluded from the analyses, resulting in a final sample of 414 patients for the main analyses. The number of observations varied across statistical models due to missing data for specific variables.

The OBESEPI study is registered at ClinicalTrials.gov (identifier: NCT02663388). The study protocol was approved by the Ethics Committee of Nancy University Hospital (CPP Est III, number: 2015-A01175-44).

### Assessment of psychological factors

All patients underwent a standardized preoperative pathway including a semi-structured psychiatric interview performed by the same psychiatrist. The interview assessed mood, anxiety, psychotic, addictive, eating, and personality disorders according to DSM-5 criteria.[17] Eating-related disturbances were assessed using a structured clinical interview based on DSM-5 criteria.[17] Diagnosed eating disorders included binge eating disorder (BED), night eating syndrome (NES),and anorexia. In addition, non-diagnostic but clinically relevant eating behaviors, such as hyperphagia and subsyndromic compulsive eating, were recorded. For the analysis, these conditions were grouped under the term “eating-related disorders and behaviors” (EDs), acknowledging the heterogeneity of this category. Psychiatric conditions included depression, anxiety disorders, bipolar disorder, addiction and intellectual disability. History of violence was assessed based on clinical reports and categorized into the following subtypes: psychological, physical, sexual, emotional neglect, and indirect violence.

### Weight trajectory outcomes

BMI was recorded at baseline (initiation of care-BMIP1), on the day of surgery (BMIP2), and at 2, 6, 12, 18, 24, 36, 48, and 60 months after surgery. (Figure 2). We selected BMI as a measure of adiposity as it is i) independent of height; ii) easy to use to calculate BMI change / delta values; iii) compatible with longitudinal series data analysis[18]. Weight regain was defined as an increase of at least 0.5 kg/m^2^ in BMI after the nadir, a threshold chosen to capture clinically meaningful early relapse in weight trajectory. Weight evolution after bariatric surgery was assessed using multiple complementary outcomes to describe weight variation:

- **Preoperative BMI change (BMIP1P2):** difference in BMI between the start of follow-up and the time of surgery
- **Maximal BMI loss (**Δ**BMImax):** maximum reduction in BMI observed during postoperative follow-up
- **Final BMI change (dBMIdf):** difference between BMI at surgery and BMI at the last available follow-up
- **Weight regain (BMIR):** defined as an increase of at least 0.5 kg/m^2^ in BMI after initial weight loss (binary outcome)
- **Magnitude of weight regain (dBMIR):** quantitative measure of BMI increase after nadir

**Figure 1:**
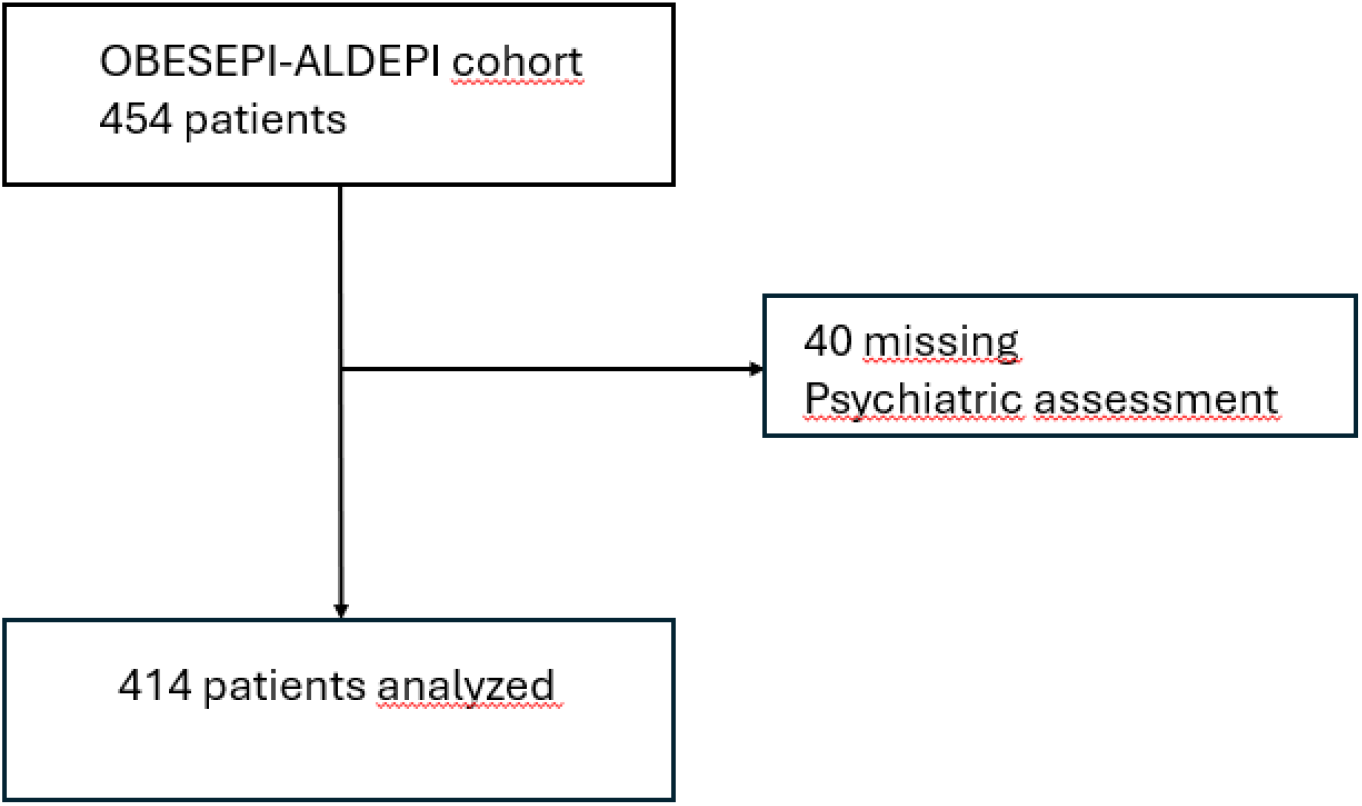
Flow-chart.

**Figure 2:**
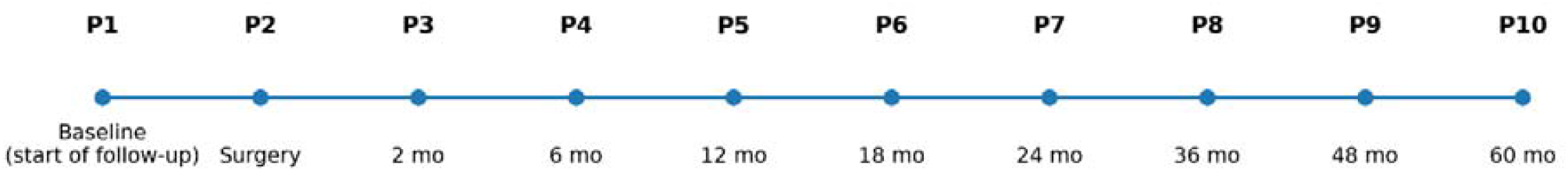
Weight measurement times.

The “L, U, D, and Y curve” profiles were designed based on i) the visual inspection of the BMI curve profiles; ii) previous validation of these profiles in longitudinal population-based and interventional designs [19, 20].Classification was performed independently by two reviewers, and based on visual inspection of individual BMI curves over time.

- **L-shaped:** initial weight loss followed by stabilization
- **U-shaped:** initial weight loss followed by sustained weight regain
- **D-shaped:** continuous decrease in BMI throughout follow-up
- **Y-shaped:** fluctuating trajectory with alternating phases of weight loss and regain

### Statistical analysis

Continuous variables are presented as mean ± standard deviation, and categorical variables as frequencies and percentages. Univariate analyses were performed using Welch’s t-test for continuous variables and Fisher’s exact test for categorical variables.

Multivariable analyses were conducted using:

- linear regression models for continuous outcomes (ΔBMImax, dBMIdf, dBMIR) adjusted for age and sex.
- logistic regression models for binary outcomes (BMIR)
- multinomial logistic regression for trajectory patterns

All models included eating disorders, psychiatric conditions, and history of violence as main independent variables. Analyses were performed using a complete case approach.

## Results

### Study population

Patients underwent four different types of bariatric surgery: laparoscopic Roux-en-Y gastric bypass (85.40%), laparoscopic sleeve gastrectomy (11.43%), duodenal switch procedure (1.90%), and laparoscopic adjustable gastric banding (1.27%). The sample was predominantly female (77%); mean age was 43.7 years, and mean BMI at the beginning of follow-up (BMIP1) was 45.85 kg/m^2^. The number of observations varied across analyses due to missing data for specific variables. Detailed sample sizes for each analysis are reported in the corresponding tables.

Among the study population:

- 362 patients (87.4%) had a history of eating disorders (EDs)
- 60 patients (14.49%) had at least one psychiatric condition
- 302 patients (72.9%) had a history of violence (Table1)

### Baseline characteristics

Baseline BMI did not differ significantly according to the presence of EDs, psychiatric conditions, or history of violence (all p > 0.30), suggesting comparable obesity severity across groups (Table 2). No significant association was observed between psychological factors and preoperative BMI variation (BMIP1P2).

**Table 1.** Characteristics of the study population.

| Variable | N (%) | Missing n (%) |
| --- | --- | --- |
| **Total sample** | 454 (100) |  |
| Eating disorders (ED) | 399 (87.8) | 40 (8.8) |
| • Binge eating disorder | 246 (54.1) | — |
| • Hyperphagia | 80 (17.6) | — |
| • Night eating syndrome | 55 (12.1) | — |
| • Anorexia | 8 (1.76) | — |
| Psychiatric conditions | 60 (13.2) | 40 (8.8) |
| • Depression | 28 (6.17) | — |
| • Anxiety | 13 (2.86) | — |
| • Bipolar disorder | 5 (1.1) | — |
| • Addiction | 12 (2.64) | — |
| • Intellectual disability | 2 (0.44) | — |
| History of violence | 302 (66.5) | 40 (8.8) |
| • Physical violence | 75 (21.2) | — |
| • Psychological violence | 74 (16.3) | – |
| • Sexual violence | 54 (11.89) | – |
| • Emotional neglect | 281 (53.08) | – |
| • Indirect violence | 73 (16.08) | – |

**Table 2:**
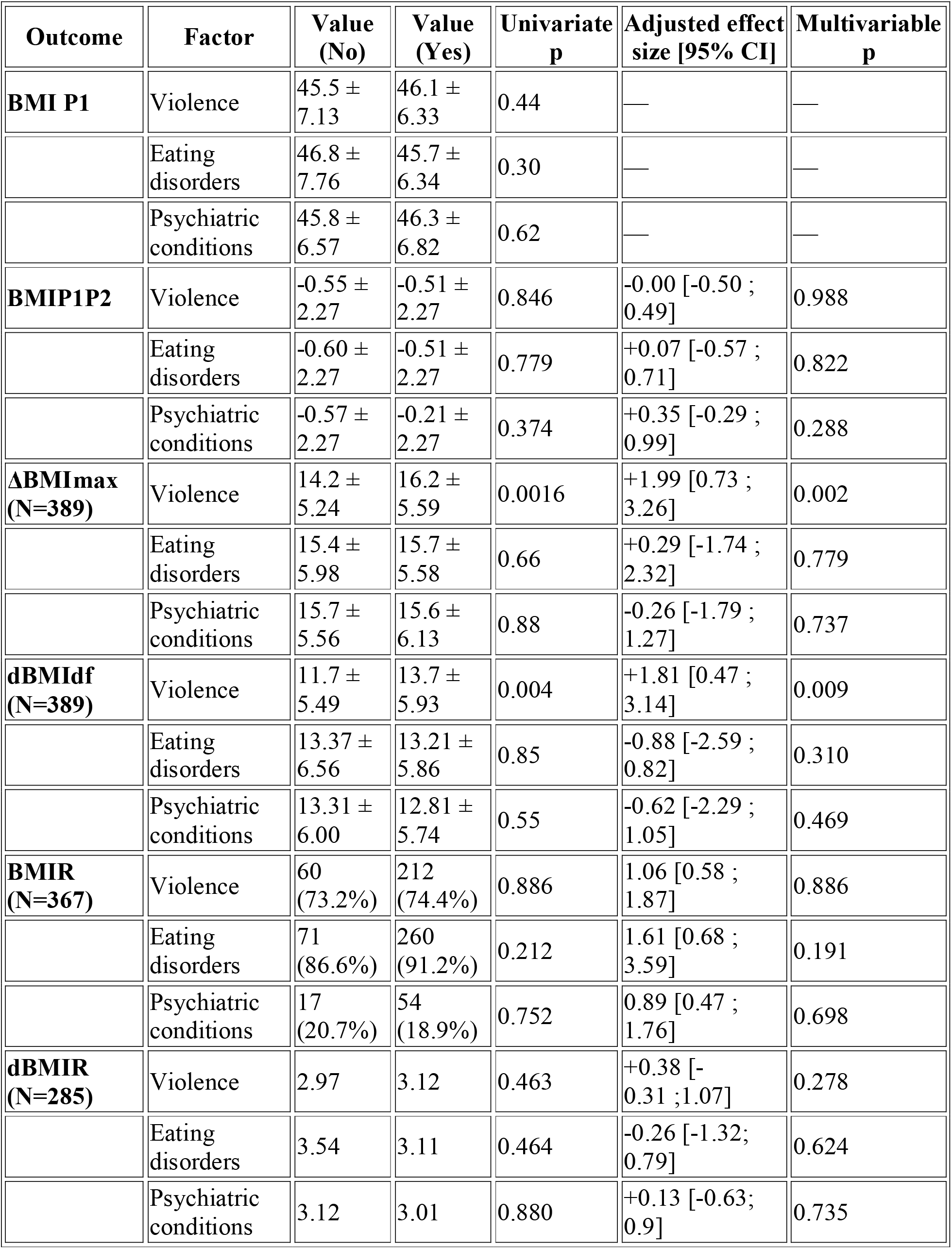
Univariate and multivariate analysis for psychiatric outcomes and weight outcomes. Continuous variables are expressed as mean ± SD. Binary outcome (BMIR) is expressed as percentages. Univariate analyses were performed using Welch’s t-test or Fisher’s exact test. Multivariable analyses were performed using linear or logistic regression models. β coefficients are presented for continuous outcomes and odds ratios (OR) for binary outcomes (BMIR). Models were adjusted for relevant age and sex

| Outcome | Factor | Value (No) | Value (Yes) | Univariate p | Adjusted effect size [95% CI] | Multivariable p |
| --- | --- | --- | --- | --- | --- | --- |
| **BMI P1** | Violence | 45.5 ± 7.13 | 46.1 ± 6.33 | 0.44 | — | — |
|  | Eating disorders | 46.8 ± 7.76 | 45.7 ± 6.34 | 0.30 | — | — |
|  | Psychiatric conditions | 45.8 ± 6.57 | 46.3 ± 6.82 | 0.62 | — | — |
| **BMIP1P2** | Violence | -0.55 ± 2.27 | -0.51 ± 2.27 | 0.846 | -0.00 [-0.50 ; 0.49] | 0.988 |
|  | Eating disorders | -0.60 ± 2.27 | -0.51 ± 2.27 | 0.779 | +0.07 [-0.57 ; 0.71] | 0.822 |
|  | Psychiatric conditions | -0.57 ± 2.27 | -0.21 ± 2.27 | 0.374 | +0.35 [-0.29 ; 0.99] | 0.288 |
| **ΔBMI<sub>max</sub> (N=389)** | Violence | 14.2 ± 5.24 | 16.2 ± 5.59 | 0.0016 | +1.99 [0.73 ; 3.26] | 0.002 |
|  | Eating disorders | 15.4 ± 5.98 | 15.7 ± 5.58 | 0.66 | +0.29 [-1.74 ; 2.32] | 0.779 |
|  | Psychiatric conditions | 15.7 ± 5.56 | 15.6 ± 6.13 | 0.88 | -0.26 [-1.79 ; 1.27] | 0.737 |
| **ΔBMI<sub>df</sub> (N=389)** | Violence | 11.7 ± 5.49 | 13.7 ± 5.93 | 0.004 | +1.81 [0.47 ; 3.14] | 0.009 |
|  | Eating disorders | 13.37 ± 6.56 | 13.21 ± 5.86 | 0.85 | -0.88 [-2.59 ; 0.82] | 0.310 |
|  | Psychiatric conditions | 13.31 ± 6.00 | 12.81 ± 5.74 | 0.55 | -0.62 [-2.29 ; 1.05] | 0.469 |
| **BMIR (N=367)** | Violence | 60 (73.2%) | 212 (74.4%) | 0.886 | 1.06 [0.58 ; 1.87] | 0.886 |
|  | Eating disorders | 71 (86.6%) | 260 (91.2%) | 0.212 | 1.61 [0.68 ; 3.59] | 0.191 |
|  | Psychiatric conditions | 17 (20.7%) | 54 (18.9%) | 0.752 | 0.89 [0.47 ; 1.76] | 0.698 |
| **ΔBMIR (N=285)** | Violence | 2.97 | 3.12 | 0.463 | +0.38 [-0.31 ; 1.07] | 0.278 |
|  | Eating disorders | 3.54 | 3.11 | 0.464 | -0.26 [-1.32 ; 0.79] | 0.624 |
|  | Psychiatric conditions | 3.12 | 3.01 | 0.880 | +0.13 [-0.63 ; 0.9] | 0.735 |

### Weight loss outcomes

Regarding the variables dBMImax and dBMIdf, 389 patients could be analysed due to missing BMI data in the post-operative follow-up. A history of violence was significantly associated with greater maximal BMI loss (ΔBMImax: 16.2 ± 5.6 vs 14.2 ± 5.2, p = 0.0016). This association remained significant in multivariable analysis (β = 1.99, 95% CI 0.73–3.26; p= 0.002). Similarly, patients with a history of violence had greater BMI reduction at final follow-up (dBMIdf: 13.7 ± 5.9 vs 11.7 ± 5.5, p = 0.004), which remained significant after adjustment (β = 1.81, 95% CI 0.47–3.14; p = 0.009).(Table2) No significant associations were observed between EDs or psychiatric conditions and weight loss outcomes.

### Weight regain

Complete data were available for 367 patients; 285 in the “regain group” and 82 in the group without regain. In univariate analysis, No significant association was found between the ‘weight regain’ status and exposure to violence (p=0.886); the presence of a psychiatric disorder (p=0.752); or the presence of eating disorders (p=0.212).A multivariate logistic regression analysis was conducted to assess the factors associated with BMIR status; in this analysis, adjustments were made for age and sex. None of the variables studied was significantly associated with BMIR status (Table 2).

The variable dBMIR corresponds to the extent of weight regain. The analysis included only the 285 patients who had regained weight and for whom complete data were available. The magnitude of weight regain (dBMIR) was not significantly associated with any psychological factors. (Table 2). Description of the BMIregain group is available in Table 3.

**Table 3:** Characteristics of patients with weight regain.Data are presented as counts (n) and percentages (%), calculated among patients with weight regain (BMIR = 1). Some patients presented multiple subtypes; therefore, percentages may exceed 100%

| Domain | Variable | n (%) |
| --- | --- | --- |
| **Global** | Violence | 212 (74.4%) |
|  | Eating disorders (TCAA) | 260 (91.2%) |
|  | Psychiatric disorders (TPA) | 54 (18.9%) |
| **Violence** | Physical | 61 (21.4%) |
|  | Psychological | 47 (16.5%) |
|  | Sexual | 37 (13.0%) |
|  | Emotional neglect | 202 (70.9%) |
|  | Indirect violence | 53 (18.7%) |
| **Eating disorders** | BED | 226 (75.3%) |
|  | Hyperphagia | 50 (16.7%) |
|  | NES | 40 (13.3%) |
|  | Anorexia | 5 (1.7%) |
|  | Bulimia | 6 (2.0%) |
| **Psychiatric disorders** | Depression | 19 (6.3%) |
|  | Addiction | 10 (3.3%) |
|  | Bipolar disorder | 2 (0.7%) |
|  | Intellectual disability | 2 (0.7%) |

### Trajectory patterns

The BMI shape profiles (L, U, D and Y) used in this analysis are described in the ‘Methods’ section. Table 4 shows the distribution of the curves according to the studied conditions. A univariate analysis was conducted to examine the association between shape type and the three studied factors. (without distinguishing by subtype). Comparisons were carried out using Pearson’s chi-squared test. Where the conditions for its application were not met, a Fisher’s exact test was performed. No statistically significant association was observed between the shapes of the weight curves and the three studied factors. Analysis of the distributions did not reveal any significant difference in the distribution of trajectory profiles between the groups. (Table 4).

**Table 4:** Distribution of BMI shape according to psychiatric factors. The results are expressed as numbers (n) and percentages (%).

| Shape | ED<br>absent<br>n (%) | ED<br>présent<br>n (%) | p | Psychiatric<br>disorder<br>absent n<br>(%) | Psychiatric<br>disorder<br>présent n<br>(%) | p | Violence<br>absent n<br>(%) | Violence<br>présent<br>n (%) | p |
| --- | --- | --- | --- | --- | --- | --- | --- | --- | --- |
| **L** | 4<br>(11.1%) | 34<br>(10.3%) |  | 31 (10.5%) | 7 (9.9%) |  | 11<br>(11.6%) | 27<br>(10.0%) |  |
| **D** | 7<br>(19.4%) | 37<br>(11.2%) |  | 34 (11.5%) | 10 (14.1%) |  | 11<br>(11.6%) | 33<br>(12.2%) |  |
| **U** | 17<br>(47.2%) | 137<br>(41.5%) |  | 121 (40.9%) | 33 (46.5%) |  | 44<br>(46.3%) | 110<br>(40.7%) |  |
| **Y** | 8<br>(22.2%) | 123<br>(37.3%) |  | 110 (37.2%) | 21 (29.6%) |  | 29<br>(30.5%) | 102<br>(37.8%) |  |
| **p-value** |  |  | **0.206** |  |  | **0.618** |  |  | **0.612** |

In a second step, we conducted a multinomial logistic regression analysis, using the L-type curve (weight loss followed by stabilisation) as the reference weight curve. This was chosen because this type of weight curve corresponds to the ‘ideal’ trajectory following surgery, while the others may be considered pathological. The results are expressed as odds ratios (OR) with their p-values, calculated using the Wald statistical test. The analyses were performed after excluding observations with missing data (398 patients analysed). In the multinomial model, no statistically significant association was observed between the studied factors and the weight trajectories (Table 5).

**Table 5:**
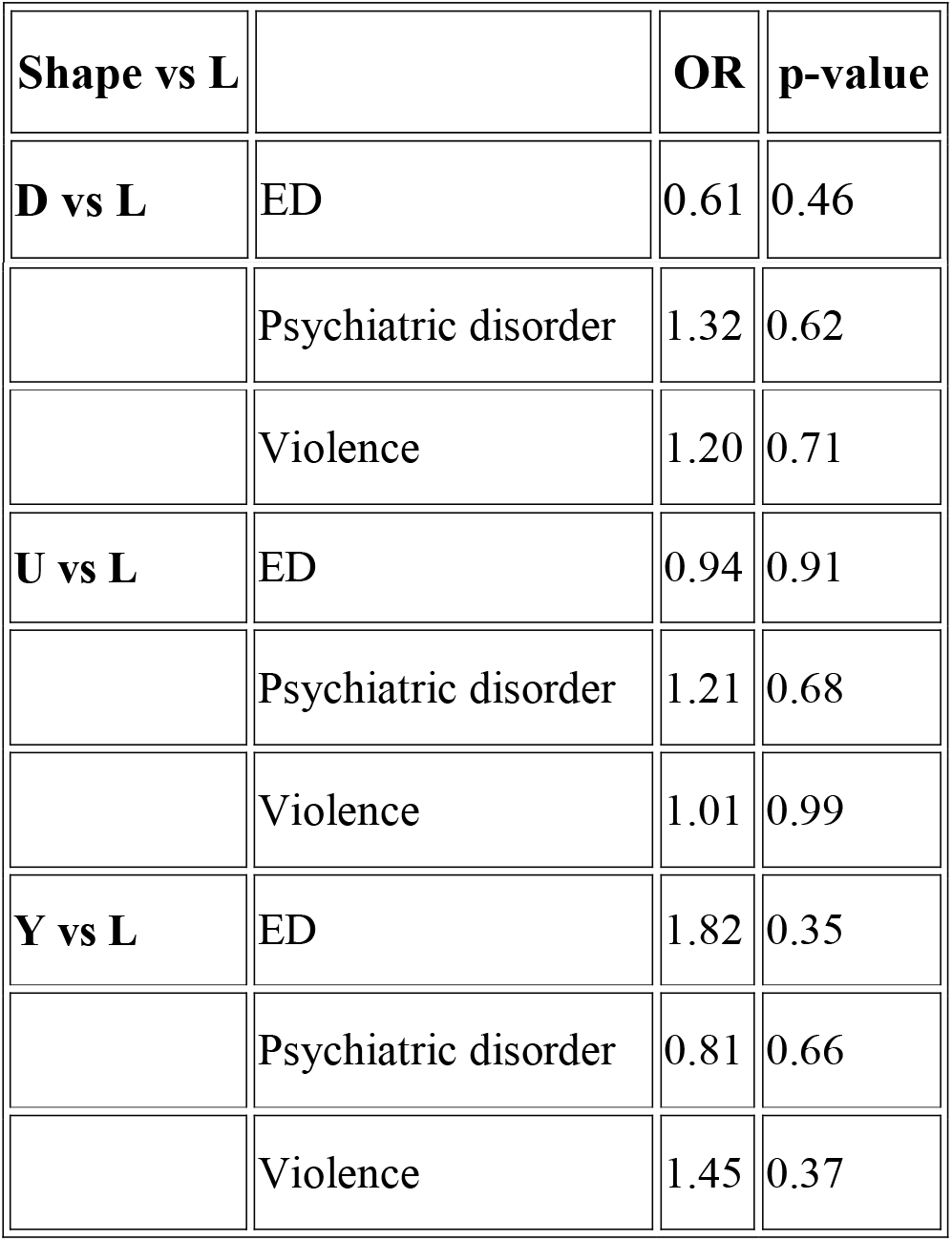
Association between weight trajectories (BMI-shape) and psychiatric factors: multinomial analysis.

### Subgroup analyses

Subgroup analysis provides insight into the various phases of weight gain. However, these results should be interpreted with caution due to the small sample sizes in some groups.

Among ED subtypes, a trend toward an association between binge eating disorder and weight regain was observed (p=0.048; IC :0.99-2.81). Among violence subtypes, only physical violence was significantly associated with weight regain (p = 0.009). These results should be interpreted cautiously due to small group size.(Table 6)

**Table 6:** Association between psychiatric subtypes and weight outcomes. Values are presented as mean ± standard deviation for continuous outcomes and percentages for BMIR. Comparisons were performed using Welch’s t-test or Fisher’s exact test. Bold values indicate statistically significant results.

|  |  |  |  |  |  |  |  |  |  |  |
| --- | --- | --- | --- | --- | --- | --- | --- | --- | --- | --- |
| **Violence** | Psychological | 16.0 vs<br>15.6 | 0.53 | 13.9 vs<br>13.0 | 0.21 | 26.8 vs<br>16.5 | 0.41<br>[0.22–<br>0.77] | **0.007** | 3.52 vs<br>3.76 | 0.55 |
|  | Physical | 15.5 vs<br>15.7 | 0.71 | 13.0 vs<br>13.3 | 0.65 | 8.5 vs<br>21.4 | 3.31[1.39–<br>7.60] | **0.005** | 3.80 vs<br>3.70 | 0.80 |
|  | Sexual | 16.5 vs<br>15.6 | 0.31 | 14.1 vs<br>13.1 | 0.37 | 13.4 vs<br>13.0 | 0.96<br>[0.45–<br>2.20] | 1.000 | 4.50 vs<br>3.61 | 0.11 |
|  | Emotional<br>neglect | 16.2 vs<br>14.6 | **0.009** | 13.7 vs<br>12.1 | **0.018** | 67.1 vs<br>70.9 | 1.19<br>[0.68–<br>2.08] | 0.498 | 3.74 vs<br>3.68 | 0.86 |
|  | Indirect<br>violence | 15.4 vs<br>15.8 | 0.59 | — | — | 15.9 vs<br>18.6 | 1.22<br>[0.61–<br>2.58] | 0.627 | 3.75 vs<br>3.71 | 0.92 |
| **Eating**<br>**disorders** | BED | 15.7 vs<br>15.7 | 0.99 | 13.2 vs<br>13.4 | 0.70 | 78.4 vs<br>68.8 | 1.67<br>[0.99–<br>2.81] | **0.048** | 3.69 vs<br>4.06 | 0.28 |
|  | Hyperphagia | 15.5 vs<br>15.7 | 0.75 | 13.2 vs<br>13.2 | 0.97 | 73.1 vs<br>76.3 | 0.87<br>[0.46–<br>1.68] | 0.643 | 3.49 vs<br>3.84 | 0.35 |
|  | NES | 15.3 vs<br>15.7 | 0.61 | 12.9 vs<br>13.3 | 0.64 | 76.9 vs<br>75.6 | 0.90<br>[0.45–<br>1.89] | 0.735 | 4.26 vs<br>3.71 | 0.22 |
|  | Anorexia | 12.7 vs<br>15.7 | 0.25 | 10.5 vs<br>13.3 | 0.35 | 83.3 vs<br>75.6 | 1.61<br>[0.18–<br>76.92] | 1.000 | 4.38 vs<br>3.78 | 0.79 |
| **Psychiatric**<br>**disorders** | Addiction | 15.5 vs<br>15.7 | 0.90 | 14.7 vs<br>13.2 | 0.46 | 75.0 vs<br>75.8 | 1.07<br>[0.27–<br>6.17] | 1.000 | 2.36 vs<br>3.83 | **0.026** |
|  | Depression | 16.3 vs<br>15.6 | 0.66 | 13.1 vs<br>13.2 | 0.92 | 73.9 vs<br>75.9 | 0.86<br>[0.33–<br>2.50] | 0.813 | 3.72 vs<br>3.79 | 0.94 |
|  | Bipolar disorder | 13.3 vs 15.7 | 0.32 | 10.8 vs 13.3 | 0.32 | 40.0 vs 76.2 | 0.21 [0.02–1.85] | 0.094 | 6.46 vs 3.77 | 0.70 |
|  | Intellectual disability | 16.8 vs 15.7 | 0.67 | 11.2 vs 13.2 | 0.67 | 100 vs 75.6 | 0.64 [0.03–38.01] | 0.566 | 5.63 vs 3.77 | 0.54 |

## Discussion

This retrospective study examined psychological characteristics of patients with severe obesity who underwent bariatric surgery. The results showed that psychiatric factors, such as psychiatric disorders, eating disorders, and exposure to violence, were not associated with initial BMI, nor with changes in BMI during the preoperative medical and psychological preparation phase. These results suggest that the groups were broadly homogeneous at the start of treatment, and that subsequent differences do not reflect heterogeneity between the groups or the influence of the studied psychiatric conditions on obesity severity. While the presence of psychiatric conditions or exposure to violence could play a role in the development of obesity [1, 2], there are currently no published studies supporting the idea that patients with these conditions suffer from more severe obesity.

In this study, exposure to violence was found to be the factor most strongly associated with weight changes following surgery, in terms of both maximum and follow-up BMI loss. No association was observed between overall exposure to violence and BMIR in the main analysis. However, subtype-specific analyses revealed contrasting associations, with physical violence independently associated with a higher risk of BMIR and psychological violence associated with a lower risk. This association with weight loss parameters was found to persist after adjustment, making it reliable. While the literature confirms the high prevalence of trauma in the obese population, it is not unanimous on this subject, with divergent results regarding the impact of such psychological trauma on weight changes following bariatric surgery. In a meta-analysis including seven studies, *Konrad et al*. did not identify an association between psychological trauma and postoperative weight loss [21]. Similarly, *Shinagawa et al*. found no difference in weight loss between patients exposed to violence and those not exposed [22]. Conversely, *Lodhia et al*. showed that patients exposed to multiple types of violence had lower weight loss one year after surgery[23]. While this result may appear to contradict our own findings, subgroup analyses revealed different associations depending on the subtype of violence. Regarding weight loss, we found that emotional neglect was associated with greater postoperative weight loss. This reinforces the need to conduct analyses by subtype of violence rather than grouping them all together as a single entity [23]. Analysis of weight regain confirms that not all types of violence should be grouped together. Indeed, an effect favouring weight regain was observed for physical violence, and the opposite effect was observed for psychological violence. This association remained significant in multivariable analysis, although it should be interpreted with caution given the sample size. These results are consistent with previous studies, which emphasise the need to distinguish between forms of trauma, their timing and their potential cumulative effect on weight variation [21, 23]. The seemingly protective nature of psychological violence must be interpreted with caution. It may stem from collinearity with other forms of violence.

Conversely, eating disorders were not associated with maximum BMI loss or BMI loss at the end of the follow-up period. This finding is consistent with the meta-analysis by Kops et al., who found no association between Binge Eating Disorder (BED) and post-operative weight loss based on 19 studies with follow-up periods of up to 60 months.[24]. In our study, a trend was observed in the association between BED and weight regain status in univariate analysis. However, this trend was not confirmed after adjusting for subtypes of violence, age, and sex. These findings suggest that BED may reflect underlying behavioural or traumatic vulnerability rather than acting as an independent determinant of weight outcomes. It also highlights the difficulty of analysing selected factors together, given their probable causal association. Indeed, eating disorders frequently result from exposure to one or more forms of violence[25–27]. In a meta-analysis, *Kops et al*. found similar results, with no association between weight loss and a preoperative diagnosis of BED[24]. *Mauro et al*. showed that the onset of eating disorders has a more significant impact on weight loss and potential weight regain, suggesting the value of psychological follow-up throughout the postoperative period to detect the emergence of de novo eating disorders.[28]

Active psychiatric disorders, analysed either globally or by diagnosis, were not significantly associated with postoperative weight variation, in terms of either weight loss or weight regain. This finding is consistent with several reviews showing that preoperative psychiatric conditions were not clearly associated with weight loss following bariatric surgery when considered holistically[28, 29]. *Sarwer et al*. argue in their study that characteristics common to all psychological diagnoses, rather than specific diagnoses, best characterise psychopathology.[29] Mood disorders and addictions, as well as binge eating and bulimia, share the characteristic of impulsivity. However, other terms are used to describe this phenomenon, such as emotional dysregulation or disinhibition (i.e. loss of control over food consumption)[29]. This supports the view that psychiatric factors, eating disorders, and exposure to violence are deeply intertwined, the impact of which is difficult to identify independently.[29] It also suggests that the medical framework for making these diagnoses is likely too rigid. It also suggests the value of conducting analyses that consider the cumulative effect of psychological conditions present at the time of surgery.

Another important contribution of this study is its distinction between the onset and extent of weight regain. Physical violence was the only factor associated with weight regain in subtype analyses, showing a tendency towards an association with BED. These results are consistent with those of *Sarwer et al*., who identified BED and depressive disorders as being associated with weight regain. [29]We did not find an association between depressive disorders and weight regain in this study but only considered a diagnosis of depressive disorder at the time of the initial psychiatric consultation. An analysis accounting for a history of depressive disorders could help clarify their association with weight fluctuations. No association was found with the extent of weight regain. This may be due to the small sample size, as only patients who had regained weight could be included in the study. An association was identified between the extent of weight regain and the presence of an addiction. However, this result cannot be interpreted due to the small sample size. These results suggest that the studied factors may influence the onset of weight regain rather than its magnitude. This distinction is important because it indicates that the determinants of occurrence and severity are not necessarily the same. It also suggests that factors other than the psychological factors studied here influence the extent of weight regain.

Regarding weight trajectory profiles, none of the analyses (univariate and multinomial regression) revealed a significant association with eating disorders, psychiatric disorders, or a history of violence. These results imply that the psychiatric factors investigated do not significantly affect the overall shape of the weight curve, despite potentially being linked to events, such as weight regain. This may also reflect a lack of statistical power in certain trajectory categories, particularly for the less frequent profiles. To date, studies examining postoperative weight changes have focused on final weight outcomes rather than dynamic curve profiles, which limits possible comparisons with the literature [28, 29].

This study has several strengths. Firstly, it is based on a well-characterised cohort, with psychiatric assessments carried out by a single psychiatrist specialising in the management of patients with severe obesity, which limits heterogeneity in phenotyping. Diagnoses were made using the DSM-5, the gold standard for psychiatric diagnosis. Secondly, the study is not limited to a single outcome measure but rather explores several dimensions of weight change using parameters that characterise both post-operative weight loss and weight regain. Thirdly, the detailed analysis by subtype of violence, eating disorders, and psychiatric disorders constitutes an original contribution, providing a precise description of the pathologies within a population suffering from severe obesity.

However, several limitations must be highlighted. The study is retrospective and certain subgroups were very small, particularly those with anorexia, bulimia, bipolar disorder or intellectual disability. This severely limits the statistical power and potential generalisation of the results. Furthermore, variables were dichotomized based on the presence of at least one condition within each domain, which may have masked potential dose–response or cumulative effects. Furthermore, some patients may have presented with multiple types of EDs, psychiatric conditions or types of violence. The cumulative effect of these conditions/exposures was not investigated. In this study, we focused on the presence of eating disorders (EDs) and psychiatric conditions at the time of the initial psychiatric consultation.

However, we did not consider patients’ histories. Taking history and the point in life at which these conditions arose into account would help highlight the causal links between the various conditions. Furthermore, the composition of the ED group also warrants caution, as it combined DSM-5 eating disorder diagnoses with clinically relevant but non-diagnostic eating-related behaviours, resulting in a heterogeneous category.

Overall, our results suggest that exposure to violence, rather than active psychiatric diagnoses or eating disorders (EDs) as a whole, is the psychological factor most closely linked to weight loss following bariatric surgery. The observed association between overall exposure to violence and both maximal and final weight loss suggests that traumatic history should be better integrated into the preoperative assessment. Conversely, the absence of a robust effect of overall psychiatric disorders confirms that simple preoperative diagnostic identification is insufficient for predicting weight change. Analyses considering lifelong history are necessary to elucidate these associations. Prospective studies on large cohorts incorporating standardised assessments of trauma, eating disorders and psychiatric disorders using a cumulative approach to these conditions could improve our understanding of their effect on weight change following bariatric surgery or during medical treatment for obesity [21, 22, 28]

## Conclusion

In conclusion, this study demonstrates that weight trajectory after bariatric care is not governed by a single mechanism but reflects the interaction of distinct psychological factors across different phases. Trauma history appears to influence weight loss dynamics, whereas eating disorders showed limited association with postoperative outcomes in this study. These findings challenge a static view of psychological comorbidities and support a dynamic, trajectory-based approach to obesity management. Integrating these dimensions into clinical practice may improve patient stratification and contribute to more personalized and effective long-term care

## Data Availability

The data that support the findings of this study are available from the corresponding author upon reasonable request

## Statements and Declarations

### Author Contributions

All authors contributed to the study conception and design. Material preparation, data collection and analysis were performed by Blanche Nitting, Coralie Gaspard and Claire Nominé-Criqui. The first draft of the manuscript was written by Blanche Nitting and Claire Nominé-Criqui, and all authors commented on previous versions of the manuscript. All authors read and approved the final manuscript.

### Data Availability

The datasets generated during and/or analysed during the current study are available from the corresponding author on reasonable request.

### Ethics approval

This study was performed in line with the principles of the Declaration of Helsinki. Approval was granted by the Ethics Committee of Nancy University Hospital (CPP Est III, number: 2015-A01175-44).

### Consent to participate

Informed consent was obtained from all individual participants included in the study.

### Consent to publish

Not applicable.

